# Setting Standards to Promote Artificial Intelligence in Colon Mass Endoscopic Sampling

**DOI:** 10.1101/19008078

**Authors:** Yueping Zheng, Ruizhang Su, Wangyue Wang, Sijun Meng, Hang Xiao, Wen Zhang, Haineng Xu, Yemei Bu, Yuhuan Zhong, Yi Zhang, Hesong Qiu, Wenjuan Qin, Yongxiu Zhang, Wen Xu, Hong Chen, Changhua Zhang, Siqi Wu, Zhaofang Han, Xiaofang Zheng, Huafen Zhu, Shuisheng Wu, Wensheng Pan, Yulong He, Yiqun Hu

## Abstract

**Objective:** Artificial intelligence (AI) has undeniable values in detection, characterization, and monitoring of tumors during cancer imaging. However, major AI explorations in digestive endoscopy have not been systematically planned, and more important, most AI productions are based on Single-center Studies (ScSs). ScSs result in data scarcity, redundancy as well as island effects, which leads to some limitations in applying it on endoscopy. We investigate the disadvantages of picture processing which may effect the AI detection, and make improvements in AI detection and image recognition accuracy.

**Design:** Current investigation aggregates a total of 2,500 gastroenteroscopy samples from various hospitals in multiple regions and carries out deep learning.

**Results:** It is found that factors inconducive to AI recognition are common such as: (a) the gastrointestinal tract is not cleaned up completely; (b) shooting angle (from left to right and the top of polyp are unexposed clearly), shooting distance (too close or too far to shoot causes the lump to be unclear), shooting light (insufficient light source or overexposed light source in mass) and unstable shooting lead to poor quality of pictures.

**Conclusion:** We set standards for a multicenter cooperation involving three-level medical institutions from the provincial, municipal and county to improve the recognition accuracy as well as the diagnosis and treatment efficiency meanwhile.

## INTRODUCTION

In the year of 2018, the Global Cancer Research Center (GCRC) estimated incidence and mortality worldwide for 36 cancers in 185 countries, and it reported that incidence and mortality rates of colorectal cancer were 10.2% and 9.2%, respectively. Morbidity of colorectal cancer is the third highest and its mortality is ranked as the second^1^. It is urgent for earlier screening, prevention and treatment of colorectal cancer. Previous investigations find that nearly a quarter of existing colonic adenomas could not been detected and diagnosed during a screening colonoscopy, while the percentage is up to 40% in recent studies^2^. Missed adenomas are a consequence of multiple factors such like poor bowel preparation^3^, insufficient time for colonoscopy withdrawal^4^, lack of expertise^5^, multiple polyps. With the development of big data-based artificial intelligence (AI) technology, AI is undeniably valuable in tumor detection, characterization, and monitoring in the diagnosis of lung cancer, brain cancer, breast cancer and prostate cancer^6^. AI is being leveraged and applied for colorectal cancer as well^7–8^. A hospital in Yokohama, Japan, has successfully developed a 500-fold endoscope that integrates AI technology to identify cancerous polyps in 0.3 seconds. The reference system makes a real-time judgment on whether the polyp is good or bad, following which the clinician can make EMR or ESD on the spot. It is known that the system uses more than 60,000 tumor cell images of more than 3,000 diagnosed cases in various hospitals in Japan to established database. The automatic cancer recognition function is formed through the deep learning analysis of tumor images in the image database. AI detects abnormalities in the lesions by defining their shapes, volumes, and surface texture. It can accurately measure the size of polyps, which will provide an objective basis for the selection of polypectomy methods. It performs histopathology to assess the disease stage, or analyze spectra to indicate suspected lesions as well as monitor period, which determine the prognosis or response to treatments over time. It greatly shortens the screening time and reduces the subjectivity of the artificial judgment, which will highly improve the tumor diagnosis efficiency as well as prognosis accuracy. Furthermore, it reduces the requirement threshold of the endoscopist experience, which can decrease the misdiagnosis rate of junior doctors. It is beneficial to popularize endoscopic technique in middle and lower level hospitals.

Although AI can help clinicians quickly identify and diagnose cancers, several factors limit its broad applications. The premise of AI diagnosis is deep learning that fit data but provide little insight into the process, so reliable and high-quality inputs are mandatory. However, the accuracy of AI feature extraction is affected by the quality of pictures owing to the subjectivity of doctors’ diagnosis and the uneven distribution of medical resources in various hospitals, especially the major AI researches of digestive endoscopy have not been systematically planned, which are lack of standardised methods for data collection and most of data are based on ScSs, resulting high data heterogeneity. Above all, data scarcity and island effect is the main problem of AI applications. Then, there is a lack of the robustness and the scalability in AI diagnosis. It is urgent to set standards for specialized AI robots so as to improve the recognition accuracy as well as the diagnosis and treatment efficiency.

## RESULTS

### Diagnosis Criteria

The current investigation collects a total of 2,500 gastroenteroscopy images from different hospitals in multiple regions. It is found that the factors inconducive to AI recognition are common, for instance, the gastrointestinal tract is not completely cleaned up, and the photography quality is low. It is troublesome to objectively and consistently evaluate the AI diagnostic efficiency in the same level. At present, a multicenter cooperation has been established among the three medical institutions from provincial, municipal and county levels. In order to take advantage of this opportunity, it is suggested that a set of standards should be taken.

### The requirements of the angle of shooting

To take photos from multiple angles of the polyp, fully exposing the root of the polyp or lump, the base and top of the polyp, and to clear glandular ducts for more information. At least five pictures are mandatory, which include four from left to right and one from the top (Figure 1).

### Clarity requirements for colonic polyps images

The polyp surface should be fully cleaned and rinsed. The process needs to avoid the influence and interference of fecal water, fecal residue and air bubble, and instruct patients to do preparation strictly following the guidelines: (a) firstly, carry out fluid diet one day before the operation; (b) then take polyethylene glycol isosmotic solution 2 L to 3 L, 250 ml per 10 min, within 2 hours, 4-6 hours before the operation in order to make sure that Boston bowel preparation scale (BBPS)^9^ is above 6 points. (BBPS is shown in Figure 2.)

In order to effectively remove the bubbles produced in the intestinal preparation process, dimethyl silicone oil 120-240 mg (3-6 ml) or 30% solution 45 ml is taken with polyethylene glycol during intestinal preparation. Illustrations of incomplete intestinal preparation are provided in Figure 3.

### Requirements for shooting light

Adjust the light of shooting, avoid reflection, insufficient light or overexposed light from the observation. Examples are shown in Figure 4.

### Requirements for shooting distance

Avoid partial or inadequate exposure. Examples are shown in Figure 5.

### Requirements about photographing machines and technology

Use a colonoscopy with high definition to check up and the host to shot at first. Make sure the shooting process is stable so as to prevent the image blur under the shakiness. Examples are demonstrated in Figure 6.

We colletct some pictures for AI to recognize according to the requirements mentioned above. It is demonstrated that a set of standards is necessary for AI recongnization. Examples are shown in Figure 7.

### Number of pictures

AI Recognition is should based on more quality and quantity of picture. As much as possible, usually at least 500 or more, we need machine to export the pictures taken excepting screen shots.

## TRAINING STANDARD

### The requirements of the angle of shooting

Take photos from multiple angles of the polyp, fully exposing the root of the polyp or lump, the base and top of the polyp, and clear glandular ducts for more information.

### Clarity requirements for colonic polyps images

Make sure the polyp surface is clean, which is rinsed completely, and avoid the influence and interference of fecal water, fecal residue and air bubble.

### Requirements for shooting light

Adjust the light of shooting, avoid reflection, insufficient or overexposed light from the observation.

### Requirements for shooting distance

Avoid partial or inadequate exposure.

### Number of pictures

AI Recognition is should based on more quality and quantity of picture. As much as possible, usually at least 500 or more, we need machine to export the pictures taken excepting screen shots.

### Requirements about photographing machines and technology

Use a colonoscopy with high definition to check up and the host to shot at first, make sure process of shooting is stable to prevent the image blur under the shakiness.

## CONCLUSION

The accuracy of AI recognization to polyps and adenomas has improved by established standard including (1) shooting angle requires that left to right and the top of polyp should be exposed completely at least five pictures; (2) intestinal preparation requires that BBPS is above 6 points. (3) the light of shooting should avoid reflection, insufficient light or overexposed light from the observation. (4) requirements for shooting distance should avoid partial or inadequate exposure. (5) use a colonoscopy with high definition to make sure process of shooting is stable to prevent the image blur under the shakines. It’s necessary to set standards to promote the development of diagnosis for colon cancer. We are in the progress of calling on more institutions to establish a multicenter partnership in medical AI. We hope to have more and more experts to elucidate and discuss the application of AI in digestive endoscopy, so that the technology can be more standardized. To promote the development of this industry, AI can truly benefit both patients and doctors.

## DISCUSSION

This result shows that AI recognition to polyps and adenomas can be more accurate by establishing standards to improve the quality of pictures. It requires at least five pictures from left to right and top to expose polyps completely, so as to make sure that no surrounding lesions are missing. The top of polyp is necessary owing to the canceration rate at the top of polyp is higher according to statistics. AI recognition to polyps would be defective owing to the cover of feces and foreign matter, so it’s essential for good preparation of gastrointestinal tract and a detailed history taking. Image acquisition machines with high quality and experienced operators can improve pictures quality.In addition, the shooting light and distance is very important. Small and medium adenomas can be captured completely and recognized. While larger adenoma.should acquire different angles of image by adjusting the body position and shooting angle, because endoscope lens is limited by far image.

The study is among the first in China to set rules and regulations for AI recognition. It can be better utilized for identification and improve efficiency. It would provide the basis for digital pathology with AI and promote the development of pathologic diagnosis, gene detection, immunity diagnosis, microbial diagnosis, as well as drug efficacy test, which will benefit the precision oncology and result in personalised care plans for patients^10^.

The development of medical products needs to emphasize ethics and data security. we need to emphasize the data monitoring since errors would require experts to discuss the source. In case machine learning algorithms or robotic systems are designed to use these tools go awry, the end user (the physician or the hospital) is always responsible to determine the responsibilities. The fault is classified as product liability due to equipment failure, and there should be a clear requirement for instructions on how to use and share the data. We currently depend on the institutional review board (IRB) and the ethics committee in the institution to guide these process. Due to the fact that the checks and balances of the IRB are not available in all hospitals and around the world^11^, we may need laws to standardize the usage of personal data in health care applications, such like the General Data Protection, to ensure that companies and individuals are always aware of patient privacy^12^.

## METHODS

The medical ethics committee in Zhongshan Hospital Xiamen University approved this study (Serial Number: XMZSYYKY_ER; Acceptance No. ky2019027).

We collected 1.6 million gastroenteroscopy images for AI recognization. It is found that 500 images didn’t meet the standards by BBPS and visual observation in the random sample of 2,500 images from various hospitals in multiple regions. After strict screening, the collected images will be resized to the appropriate zoom size and RGB channel normalization, then we make segmentation and multi-labels classification of the image. Lastly, input them into the deep learning network.

Convolutional neural networks (CNNs) are the gold standard for image analysis. It takes advantage of the local structural relationships in the image and create progressively more complex abstract representations from layer to layer. In comparison with other algorithms, CNNs incorporate very little preprocessing and it learns the filters that were previously hand-engineered in more traditional algorithms. A CNN model was used in this study^13^. Overall workflow of feature extraction are shown in Figure 8.

## Data Availability

Data would be available upon requests to corresponding authors.

## Acknowledgments

We would like to thank: (1) Yonghe Zhou, Kunping HuangZi Chong Kuo and Xiang Zhou for coordinating the data in the primary hospitals; (2) Dr. Yan Lichong of Yale University in the United States providing valuable advice in the standard; and (3) Volunteer Joey Yau for images collection work.

## Author contributions

Yiqun Hu is general manager the of the project. Hesong Qiu is the project coordinator. Wensheng Pan, Sijun Meng, Yongxiu Zhang, Yueping Zheng, Ruizhang Su, Yi Zhang, Yuling Zhong, Hong Chen, Changhua Zhang, Huafen Zhu, and Yulong He have contributed in providing clinical theory and seting standards. Shuisheng Wu and Wen Xu are committed to seting standards. Shuisheng Wu and Wensheng Pan’s contributed equally in the project.Wen Zhang and Haineng Xu contribute to manuscript writing. Yemei Bu and Wenjuan Qin have organized and labeled photos. Siqi Wu, Zhaofang Han, Xiaofang Zheng have classified and integrated data.

## Competing interests

The authors declare no competing interests.

## Funding

2019 Medical Innovation Project in Fujian Province, China: Application of Deep Learning Image Analysis Technology and Gene Association in Early Colorectal Cancer (grant no. 2019-CXB-31)

## FIGURE LEGENDS

**Figure 1.** The requirements of the angle of shooting. Each picture includes before and after AI identification. **a**, The ducts of glands are clear. **b,c**, The top of lump is fully exposed. **c**, The left and right rings are all exposed clearly. **d**, The ducts of glands are not clear and the base is not clear enough owing to the interference of fecal water. **e**, To tell if a polyp or early cancer from the side view. **f**, The top of lump is incompletely shown. But this six polyps is not exposed completely.

**Figure 2.** Clarity requirements based on the BBPS. The BBPS divides the colon into the left colon (including the sigmoid colon and rectum), the transverse colon (including hepatic flexure and splenic flexure of colon), the right half colon (including the cecum, the ascending colon) he cleanliness is into 4 grades: 0 There is solid stool that could not be removed in the colon, and the mucosa could not be seen. It was found in the people who were not prepared before operation.1 The part of the mucosa in the intestinal segment is clearly shown, and some of the mucous membrane is not clearly displayed due to feces and untransparent liquid. 2 There are a small amount of feces and untransparent liquid remaining in the colon, and the mucous membrane is shown to be cleared. 3 all the mucosa showed clear, there is no fecal or untransparent liquid residue in the colon. The score is the sum of the three segment colorectum scores.

**Figure 3.** Clarity requirements for colonic polyps images. Each picture includes before and after AI identification. **a,b,d**,Polyp is partially covered with untransparent liquid. **c,d**,Thepolyp is influenced by air bubble. **f**,There are still some feces covering in the mucus; I’ve got something in sight, so we the lump is deficient. BBPS for a/b/d/e are 1 point;c and f are 3 points.

**Figure 4.** Requirements for shooting light. Each picture includes before and after AI identification. **a,b**, Insufficient light source in mass. **c,d**,Overexposed light source in mass.

**Figure 5.** Requirements for shooting distance. **a**, It’s too close that the lump to fully exposed. **b,c**, It’s so far that information about the mass is hidden in the picture, we can’t see the details of the mass.

**Figure 6.** Requirements about photographing machines and technology. **a-c**, The process of shooting is unstable,so there are some blurred image under the shakiness.

**Figure 7.** Pictures taken in accordance with this standard. There are six images taken by department of Gastroenterology, Zhongshan Hospital Xiamen University, which strictly accords with requirements above. It includes four images taken from the sides and one from the top.See the supplyment for more examples.

**Figure 8.** AI feature extraction work flow.

## Notes

### Competing Interest Statement

The authors have declared no competing interest.

### Author Declarations

All relevant ethical guidelines have been followed and any necessary IRB and/or ethics committee approvals have been obtained.

Any clinical trials involved have been registered with an ICMJE-approved registry such as ClinicalTrials.gov and the trial ID is included in the manuscript.

